# The phenotypic spectrum of terminal 6q deletions based on a large cohort derived from social media and literature: a prominent role for *DLL1*

**DOI:** 10.1101/2022.11.08.22282043

**Authors:** Aafke Engwerda, Wilhelmina S. Kerstjens-Frederikse, Nicole Corsten-Janssen, Trijnie Dijkhuizen, Conny M. A. van Ravenswaaij-Arts

## Abstract

**Background:** Terminal 6q deletions are rare, and the number of well-defined published cases is limited. Since parents of children with these aberrations often search the internet and unite via international social media platforms, these dedicated platforms may hold valuable knowledge about additional cases. The Chromosome 6 Project is a collaboration between researchers and clinicians at the University Medical Center Groningen and members of a Chromosome 6 support group on Facebook. The aim of the project is to improve the surveillance of patients with chromosome 6 aberrations and the support for their families by increasing the available information about these rare aberrations. This parent-driven research project makes use of information collected directly from parents via a multilingual online questionnaire. Here, we report our findings on 93 individuals with terminal 6q deletions and 11 individuals with interstitial 6q26q27 deletions, a cohort that includes 38 newly identified individuals.

**Results:** Using this cohort, we can identify a common terminal 6q deletion phenotype that includes microcephaly, dysplastic outer ears, hypertelorism, vision problems, abnormal eye movements, dental abnormalities, feeding problems, recurrent infections, respiratory problems, spinal cord abnormalities, abnormal vertebrae, scoliosis, joint hypermobility, brain abnormalities (ventriculomegaly/hydrocephaly, corpus callosum abnormality and cortical dysplasia), seizures, hypotonia, ataxia, torticollis, balance problems, developmental delay, sleeping problems and hyperactivity. Other frequently reported clinical characteristics are congenital heart defects, kidney problems, abnormalities of the female genitalia, spina bifida, anal abnormalities, positional foot deformities, hypertonia and self-harming behaviour. The phenotypes were comparable up to a deletion size of 7.1 Mb, and most features could be attributed to the terminally located gene *DLL1*. Larger deletions that include *QKI* (>7.1 Mb) lead to a more severe phenotype that includes additional clinical characteristics.

**Conclusions:** Terminal 6q deletions cause a common but highly variable phenotype. Most clinical characteristics can be linked to the smallest terminal 6q deletions that include the gene *DLL1* (>500 kb). Based on our findings, we provide recommendations for clinical follow-up and surveillance of individuals with terminal 6q deletions.

## Background

Terminal 6q deletions are a variable group of chromosome disorders, with the largest deletions extending from 6q25.2 to 6qter (up to 16 Mb in size) and the smallest deletions restricted to the most distal band 6q27 (as small as 390 kb in size)^1^. As these deletions are rare, there is only limited information about their effect on the clinical phenotype. This lack of knowledge impairs the ability of health professionals to guide these individuals and their families.

To date, two large reviews on terminal 6q deletions have been published. Hopkin et al. (1997) described 26 individuals with deletions in the 6q25q27 region. These individuals were diagnosed by conventional cytogenetic methods, and interstitial deletions including only the proximal part of 6q27 were also considered to be terminal^2^. Lee et al. (2011) described another 28 individuals with pure terminal 6q deletions diagnosed by conventional cytogenetic methods or array CGH, with all findings confirmed by FISH^3^. The individuals described in these two reviews presented with a variable phenotype including brain abnormalities, hydrocephalus, microcephaly, seizures, intellectual disability, hyperactivity/attention deficit hyperactivity disorder (ADHD), hypotonia, club feet, genital hypoplasia, retinal abnormalities, cleft palate, dimpling on elbows and knees and facial dysmorphisms including ear anomalies, broad nasal tip, prominent nasal bridge, epicanthic folds and short palpebral fissures^2,3^.

Whereas earlier studies mostly used conventional cytogenetic methods, detailed microarray techniques are now the routine diagnostic method. The robust and detailed microarray technique allows for reliable comparisons of cytogenetic results from all over the world. In contrast, international collection of detailed phenotypes lags behind the collection of genetic data because only a minority of rare chromosomal aberration cases are submitted to international databases like DECIPHER (https://www.deciphergenomics.org), and case reporting relies on health professionals having the time and willingness to submit information. As a result, clinical information is often incomplete.

Nowadays parents of individuals with a rare disorder often start searching the internet for more information, and these parents frequently unite on social media platforms. In 2013, we started the Chromosome 6 Project, a successful collaboration with a Chromosome 6 parent support group on Facebook^4^. The Chromosome 6 Project allowed us to study detailed clinical information for 35 newly identified individuals with terminal 6q deletions. The addition of information about 58 individuals reported in literature who were diagnosed by a high-resolution cytogenetic technique allowed us to describe the phenotype based on eight subgroups with different sizes of terminal 6q deletions. We could also describe the phenotype of interstitial 6q26 and 6q27 deletions by including three newly identified individuals and eight cases from the literature. Where relevant, we discuss the candidate genes for specific clinical characteristics.

Our large cohort of 93 individuals with terminal 6q deletions allowed us to describe the most clinically relevant phenotype and define the effect of deletion size on this phenotype. Based on this data, it appears that deletion of the gene *DLL1* (Delta-Like Canonical Notch Ligand 1, MIM*606582) plays the most prominent causal role in the terminal 6q deletion phenotype. Our clinical description also leads to recommendations for clinical follow-up and surveillance that will enable healthcare professionals to better inform families and provide the best possible guidance to individuals with terminal 6q deletions.

## Methods

To describe the terminal 6q deletion phenotype, we collected detailed genotype and phenotype data on as many individuals as possible. The recruitment of individuals via social media, collection of clinical information directly from parents (parent cohort), collection of patients extracted from the literature (literature cohort) and data analysis were performed as described previously^4^ and are outlined in short below.

### Parent cohort

Individuals were informed about the Chromosome 6 Project and approached to participate via Facebook (Chromosome 6 Facebook group), Twitter (@C6study) and our website (https://www.chromosome6.org). Patients or their legal representatives could participate by signing up via our secured website. Participants received a personal account for the online Chromosome 6 Questionnaire if there was an isolated chromosome 6 aberration and a microarray report was available. Informed consent was obtained through the questionnaire. This procedure was ethically waived by the accredited Medical Ethics Review Committee of the University Medical Center Groningen.

Genotype data was extracted from the microarray reports, which were uploaded as part of the sign-up procedure. These microarray analyses were performed in diagnostic laboratories using different platforms. The UCSC LiftOver Tool was used to convert the microarray results to GRCh37/hg19, and the UCSC genome browser was used to visualise the results (http://genome.ucsc.edu).

In the current study we focus on participants with a terminal 6q deletion: a deletion extending to the end of 6q, defined as including the most distally located gene (*PDCD2*, MIM*600866), or an interstitial deletion within the 6q26q27 region (161,000,001–171,115,067).

Phenotype information on pregnancy and birth, congenital abnormalities, relevant dysmorphic features, development, behaviour and health of the child was collected via the multilingual online Chromosome 6 Questionnaire, which was constructed with the MOLGENIS toolkit^5^. Clinical photographs and additional consent for publication were collected. Data collected from individuals in the parent cohort was submitted to the DECIPHER database (https://decipher.sanger.ac.uk) IDs 489709–489746.

### Literature cohort

Case reports involving terminal 6q deletions were collected using PubMed and the following search criteria: (deletion or monosomy) and (6q26 or 6q27 or terminal 6q). The references were then checked for additional relevant case reports. Publications reporting detailed clinical information and microarray results or comparably detailed breakpoint analyses were included. Clinical information was extracted from the reports using the items of the Chromosome 6 Questionnaire.

### Participant characteristics and genotypes

Up to September 2021, the Chromosome 6 Questionnaire was completed and submitted by 129 parents, and this included information on 38 individuals with aberrations in the region of interest for this paper. Of these 38, 35 individuals (27 females and 8 males) had a terminal 6q deletion. Another 3 individuals (2 females and 1 male) had an interstitial deletion within the 6q26q27 region. Our literature cohort comprises 58 terminal 6q deletion cases (34 females and 24 males) and 8 interstitial 6q26q27 cases (2 females and 6 males) derived from 29 published papers^1,6–33^. In total, 93 terminal 6q deletion cases and 11 interstitial 6q26q27 cases were collected. The median age (years;months) of individuals in the parent cohort was 4;6 (range 0;6–32;9) and 12;0 (range 0;0–57;0) in the literature cohort. Data on foetuses was included both in the parent cohort (1 foetus; 23 weeks gestation) and the literature cohort (7 foetuses; median 24 (range 14–30) weeks gestation). For six out of eight of these cases, it was known that the pregnancy was terminated. For the foetus included in the parent cohort, the parents gave us consent to fill out the Chromosome 6 Questionnaire based on the ultrasound and autopsy results. Since the pregnancy was terminated, not all questions could be completed.

Although we focused on isolated terminal and interstitial deletions in the 6q26q27 region, some small additional rearrangements of other chromosomes were included based on their size and gene content.

### Data analysis

Clinical and behavioural characteristics were described as being present, absent or unknown. They are presented here as present/known, where known indicates knowledge of presence or absence. The developmental delay (intelligent quotient (IQ)) was categorised as normal (>85), borderline (70– 85), mild (50–70), moderate (30–50) or severe (<30) for individuals above the age of 2 years. This was based on formal IQ tests or, if these were not available, the mean of the developmental quotients for the milestones ‘walking independently’ and ‘using two-word sentences’. The developmental quotients were calculated as the 50^th^ centile of the population age of achievement for that milestone divided by the age of that achievement by the participant times 100. The 50^th^ centiles of the population age of achievement for the milestones ‘walking independently’ and ‘using two-word sentences’ are 12^34^ and 21 months^35^, respectively. For specific milestones, the age and range of achievement were presented, and we used a Mann-Whitney U-test to calculate whether there was a significant difference in the age of milestone achievement between individuals with smaller (those not including the gene *QKI*) and larger (those including *QKI*) terminal deletions.

The clinical characteristics of all terminal deletions were described as one group, but we also describe subgroups made to provide information on differently sized deletions and interstitial deletions separately. The terminal 6q deletion subgroups were based on the number of genes involved with a predicted haploinsufficiency (HI) effect. To determine whether genes had a predicted HI-effect, we used HI- and loss-of-function intolerance (pLI) scores as described previously^4^ (https://www.deciphergenomics.org)^36^ (http://exac.broadinstitute.org)^37^, see Table S1. Genes with a HI score <10% and/or a pLI score ≥0.9 were considered HI-genes. The terminal 6q26q27 region contains eight such HI-genes: *TBP, PSMB1, DLL1, AFDN, PDE10A, QKI, PRKN* and *MAP3K4*. This allocation resulted in eight terminal 6q deletion subgroups: T-PSMB1 (including *PSMB1* and *TBP*), T- DLL1, T-AFDN, T-PDE10A, T-QKI, T-PRKN, T-MAP3K4 and a residual group (T-R) that includes deletions extending beyond 6q26 and including the HI-gene *IGF2R* (Figure 1). Two subgroups of interstitial deletions were described: interstitial deletions of 6q26 (I-6q26) and interstitial deletions of 6q27 (I- 6q27) (Figure 1). The phenotypes of individuals with a terminal 6q deletion due to a ring chromosome were compared to those with a simple terminal deletion. We also investigated how often breakpoints in the fragile site *FRAE6* (6q26) were present in terminal 6q deletions in order to explore its involvement in breakpoint occurrence.

**Figure 1.**
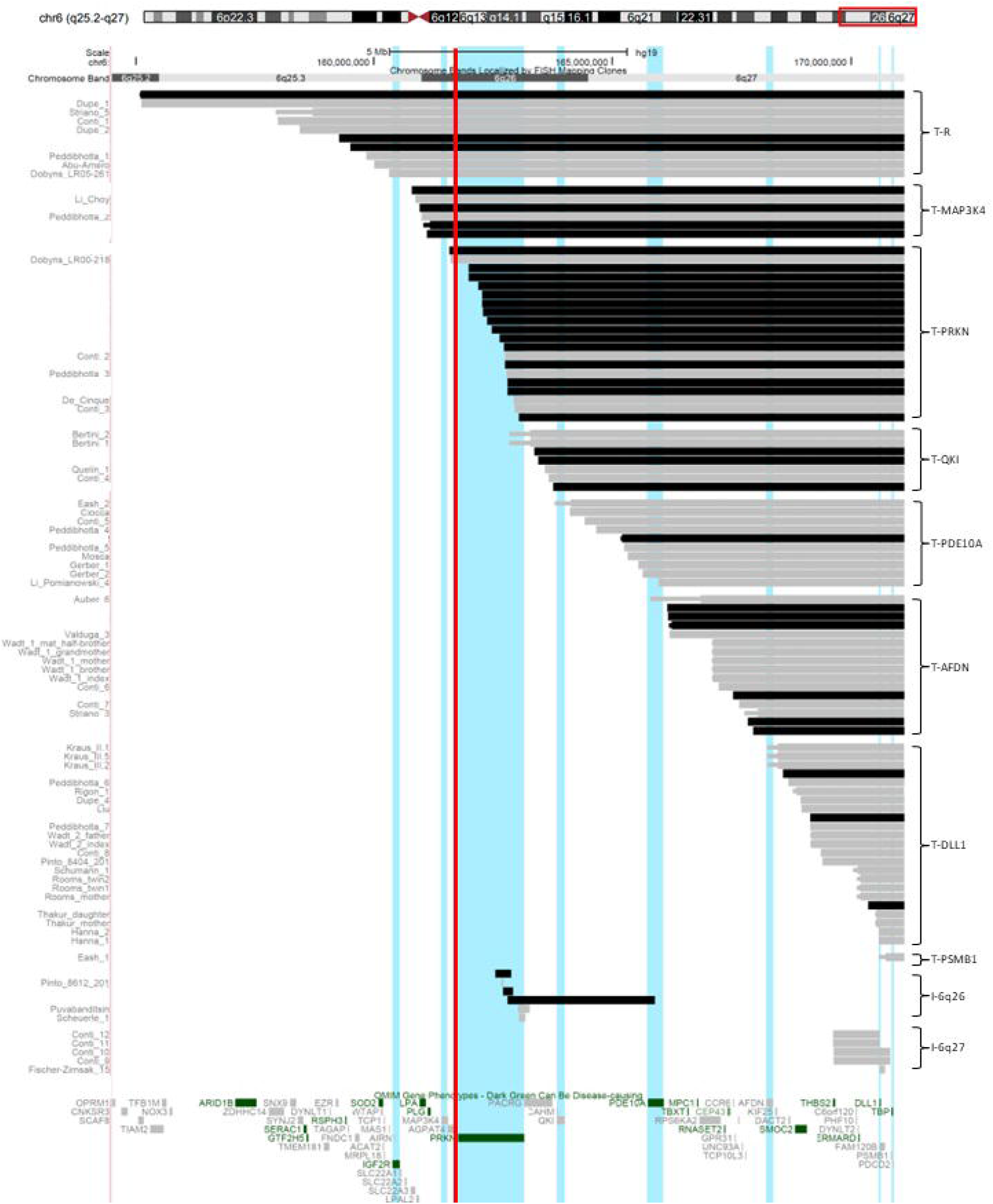
Overview of all terminal 6q deletions and interstitial 6q26q27 deletions. Deletions from our parent cohort (black bars) and literature cohort (grey bars) are shown, and their minimum (thick bar) and maximum (thin bar) deletion size are visualised when available. The terminal deletions are divided into eight subgroups (shown on the right hand side) based on the most proximal gene with a predicted haploinsufficiency effect (HI-gene) (vertical light blue lines) involved in the minimum deletion size. T-PSMB1: terminal deletions only including *PSMB1* and *TBP* (breakpoint distal to 170.6 Mb), T-DLL1: terminal deletions including *DLL1* (breakpoint between 168.4 and 170.6 Mb), T-AFDN: terminal deletions including *AFDN* (166.1–168.4 Mb), T-PDE10A: terminal deletions including *PDE10A* (164.0–166.1 Mb), T-QKI: terminal deletions including *QKI* (163.1–164.0 Mb), T-PRKN: terminal deletions including *PRKN* (161.5–163.1 Mb), T-MAP3K4: terminal deletions including *MAP3K4* (160.5–161.5 Mb) and T-R (residual group): terminal deletions including *IGF2R* and larger (breakpoint proximal to 160.5 Mb). Interstitial deletions are divided in two subgroups based on their cytogenetic location: I-6q26: interstitial deletions (mostly) located on chromosome band 6q26 (161–164.5 Mb), I- 6q27: deletions located on chromosome band 6q27 (164.5–171.1 Mb). The deletions are visualised using the UCSC genome browser (https://genome.ucsc.edu). The vertical red line marks the location of the common fragile site FRA6E. The literature cases were derived from 29 reports^1,6–33^.

## Results

All individuals were assigned to one of the ten subgroups based on the number of HI-genes present in their terminal 6q deletion or the cytogenetic location of the interstitial deletion. Figure 1 visualises the deletions, subgroups and HI-genes. The main genotype characteristics of the subgroups are summarised in Table 1. The phenotype information is summarised in Table 2 (see supplementary Table S2, for details). Information on development is given in Table 3 and visualised in Figure 2 and Figure S1. Below, we provide a phenotype description for the whole group of terminal 6q deletions, with subgroups highlighted when specific clinical characteristics were only seen in the subgroups with larger terminal deletions. Descriptions of the most prominent characteristics in the interstitial 6q26q27 deletions are given separately. In the Discussion, we briefly consider the role of the deleted genes in the phenotype, if applicable.

**Table 1.** Genotype characteristics of terminal 6q and interstitial 6q26q27 deletions

| Subgroup | Cohort<br>parent/<br>literature | No. of<br>foetuses | Age median*<br>(range) years;<br>months | Deletion size median<br>(range) Mb | No. of OMIM<br>genes median<br>(range) | No. of HI genes<br>median (range) |
| --- | --- | --- | --- | --- | --- | --- |
| Total terminal (n=93) | 35/58 | 8 | 7;5 (0;0–57;0) | 6.45 (0.39–16.00) | 25 (3–62) | 4 (1–14) |
| Parent cohort | 35/0 | 1 | 4;6 (0;6–32;9) | 8.39 (0.76–16.00) | 29 (5–62) | 6 (2–14) |
| Literature cohort | 0/58 | 7 | 12;0 (0;0–57;0) | 4.14 (0.39–15.99) | 22 (3–62) | 3 (1–14) |
| T-PSMB1 (n=1) | 0/1 | 0 | 8;0 | 0.39 | 3 | 1 |
| T-DLL1 (n=23) | 3/20 | 2 | 17;0 (0;0–49;0) | 1.95 (0.53–2.66) | 10 (5–12) | 2 |
| T-AFDN (n=16) | 6/10 | 2 | 6;6 (0;6–57;0) | 4.01 (3.07–4.97) | 22 (14–24) | 3 |
| T-PDE10A (n=10) | 1/9 | 0 | 3;10 (0;1–25;0) | 5.89 (5.16–7.00) | 25 | 4 |
| T-QKI (n=7) | 3/4 | 0 | 7;11 (0;8–25;0) | 7.68 (7.37–7.84) | 28 (27–28) | 5 |
| T-PRKN (n=20) | 15/5 | 0 | 5;0 (0;4–32;9) | 8.57 (8.09–9.53) | 29 (29–30) | 6 |
| T-MAP3K4 (n=6) | 4/2 | 2 | 6;3 (4;10–14;9) | 10.14 (9.94–10.31) | 33 (31–35) | 7 |
| T-R (n=10) | 3/7 | 2 | 8;0 (0;6–37;0) | 12.12 (10.79–16.00) | 51.5 (40–62) | 11.5 (8–14) |
| Total Interstitial<br>(n=11) | 3/8 | 0 | 6;0 (0;1–43;0) | 0.33 (0.04–3.08) | 2 (1–7) | 1 (1–3) |
| Parent cohort | 3/0 | 0 | 6;0 (2;2–15;4) | 0.33 (0.21–3.08) | 1 (1–5) | 1 (1–3) |
| Literature cohort | 0/8 | 0 | 10;2 (0;1–43;0) | 0.61 (0.04–1.18) | 4.5 (1–7) | 1 |
| I-6q26 (n=6) | 3/3 | 0 | 10;2 (0;1–19;0) | 0.23 (0.04–3.08) | 1.5 (1–5) | 1 (1–3) |
| I-6q27 (n=5) | 0/5 | 0 | 6;0 (1;6–43;0) | 0.97 (0.12–1.18) | 7 (2–7) | 1 |

\*If known, foetuses were excluded from the age calculations

**Table 2.** Overview of most prominent characteristics seen in individuals with terminal 6q deletions and interstitial 6q26q27 deletions

| Group* | T-PSMB1<br>(n=1) | T-DLL1<br>(n=23) | T-AFDN<br>(n=16) | T-PDE10A<br>(n=10) | T-QKI<br>(n=7) | T-PRKN<br>(n=20) | T-MAP3K4<br>(n=6) | T-R<br>(n=10) | Total<br>terminal<br>(n=93) | I-6q27<br>(n=5) | I-6q26<br>(n=6) |
| --- | --- | --- | --- | --- | --- | --- | --- | --- | --- | --- | --- |
| Characteristics |  |  |  |  |  |  |  |  |  |  |  |
| Sex (female/male) | male | 14/9 | 9/7 | 9/1 | 4/3 | 10/10 | 6/0 | 9/1 | 61/32 | 2/3 | 2/4 |
| Prolonged neonatal jaundice | n.a. | 0/3 | 0/5 | 1/3 | 1/4 | 6/15 | 1/4 | 0/4 | 9/38 | 0/0 | 0/5 |
| Low birth weight (<p10) | + | 0/5 | 3/9 | 0/5 | 1/6 | 5/15 | 1/4 | 2/3 | 13/48 | 0/1 | 1/4 |
| Short stature (<p10) | + | 1/7 | 1/9 | 2/7 | 0/5 | 6/15 | 1/3 | 3/3 | 15/50 | 0/1 | 0/2 |
| Head circumference<br><p10/normal/>p90 | <p10 | 5/4/2 | 2/5/1 | 4/2/2 | 5/1/1 | 13/3/0 | 1/1/0 | 3/0/1 | 34/16/7 | 1/0/0 | 0/2/0 |
| Plagiocephaly | n.a. | 2/4 | 2/6 | 0/1 | 2/3 | 7/13 | 2/4 | 1/3 | 16/34 | 0/0 | 1/4 |
| Hypertelorism | n.a. | 3/8 | 4/8 | 3/4 | 4/6 | 8/18 | 2/3 | 4/8 | 28/55 | 1/2 | 1/3 |
| Vision problems | n.a. | 2/4 | 5/6 | 1/5 | 4/5 | 10/16 | 3/4 | 5/5 | 30/45 | 0/0 | 2/4 |
| Mild vision problems | n.a. | 1/2 | 3/5 | 0/1 | 3/4 | 6/10 | 2/3 | 1/5 | 16/30 | 0/0 | 1/2 |
| Hypermetropia | n.a. | 0/4 | 2/6 | 0/4 | 4/5 | 5/17 | 3/4 | 0/4 | 14/44 | 0/0 | 2/4 |
| Nystagmus | - | 0/5 | 2/8 | 0/5 | 2/6 | 3/18 | 2/4 | 2/6 | 11/53 | 0/1 | 1/4 |
| Strabismus | + | 1/5 | 4/8 | 1/5 | 3/6 | 8/18 | 2/4 | 4/6 | 24/53 | 1/1 | 0/4 |
| Abnormal eye morphology | n.a. | 0/3 | 1/5 | 0/4 | 0/4 | 7/16 | 0/4 | 4/6 | 12/42 | 0/0 | 0/3 |
| Dysplastic outer ear | + | 5/7 | 3/8 | 5/6 | 3/5 | 7/15 | 0/4 | 3/7 | 27/53 | 1/2 | 0/3 |
| Cleft lip and/or palate | n.a. | 0/9 | 0/6 | 0/5 | 0/5 | 1/13 | 0/4 | 2/7 | 3/49 | 0/1 | 0/2 |
| Dental problems | n.a. | 2/3 | 1/4 | 1/1 | 2/5 | 7/15 | 0/3 | 0/3 | 13/34 | 0/1 | 0/3 |
| Feeding difficulties | n.a. | 3/3 | 3/6 | 2/2 | 2/5 | 13/16 | 3/4 | 2/3 | 28/39 | 0/0 | 2/3 |
| Constipation | n.a. | 0/3 | 2/7 | 1/1 | 1/3 | 5/16 | 2/4 | 1/3 | 12/37 | 0/0 | 1/3 |
| Anal abnormality | n.a. | 0/3 | 0/6 | 0/1 | 1/3 | 4/14 | 0/4 | 0/3 | 5/34 | 0/0 | 0/3 |
| Respiratory problems | n.a. | 1/3 | 2/7 | 1/2 | 0/3 | 4/15 | 0/4 | 2/4 | 10/38 | 0/0 | 1/4 |
| Recurrent infections | n.a. | 2/4 | 1/5 | 0/1 | 2/3 | 5/11 | 2/4 | 1/3 | 13/31 | 0/0 | 1/3 |
| Congenital heart defect | n.a. | 0/3 | 1/6 | 3/5 | 0/6 | 6/17 | 0/4 | 2/4 | 12/45 | 0/0 | 1/5 |
| Kidney abnormality | n.a. | 0/3 | 2/7 | 0/3 | 1/3 | 2/16 | 1/4 | 1/4 | 7/40 | 1/1 | 1/4 |
| Abnormal genitals in females | n.a. | 0/1 | 1/6 | 0/1 | 1/3 | 3/10 | 0/4 | 1/3 | 6/28 | 0/0 | 0/2 |
| Sacral dimple | n.a. | 0/4 | 3/5 | 1/2 | 3/3 | 11/14 | 2/4 | 3/3 | 23/35 | 0/0 | 1/3 |
| Spina bifida | n.a. | 0/3 | 0/6 | 0/2 | 0/3 | 3/12 | 0/4 | 2/5 | 5/35 | 0/0 | 1/4 |
| Spinal cord abnormality | n.a. | 1/4 | 0/4 | 2/5 | 3/3 | 6/14 | 1/4 | 2/4 | 15/38 | 0/0 | 0/3 |
| Abnormal vertebral morphology | + | 1/5 | 1/7 | 0/2 | 1/3 | 1/11 | 2/5 | 1/4 | 8/38 | 0/0 | 0/3 |
| Scoliosis | n.a. | 1/6 | 0/6 | 0/2 | 2/4 | 3/14 | 0/4 | 0/3 | 6/39 | 0/0 | 1/5 |
| Hip dysplasia | n.a. | 0/1 | 0/1 | 1/2 | 0/1 | 5/10 | 0/1 | 0/1 | 6/17 | 0/0 | 0/1 |
| Hypermobility of the joints | n.a. | 3/4 | 9/11 | 2/3 | 3/5 | 8/17 | 4/4 | 3/6 | 32/50 | 1/1 | 0/3 |
| Pes planus | n.a. | 0/4 | 1/7 | 0/3 | 2/5 | 2/15 | 0/4 | 0/5 | 5/43 | 0/0 | 0/5 |
| Positional foot deformity | n.a. | 0/4 | 1/7 | 2/3 | 1/5 | 1/15 | 0/4 | 3/5 | 8/43 | 0/0 | 1/5 |
| Capillary hemangioma | n.a. | 0/5 | 1/5 | 0/3 | 0/4 | 1/13 | 1/4 | 0/3 | 3/37 | 0/1 | 0/4 |
| Hypopigmentation of the skin | n.a. | 0/5 | 0/5 | 1/3 | 0/4 | 1/13 | 1/4 | 0/3 | 3/37 | 0/1 | 0/4 |
| Brain abnormalities on MRI or CT | + | 14/17 | 11/12 | 9/10 | 6/7 | 17/18 | 5/5 | 9/9 | 72/79 | 4/5 | 2/2 |
| Ventriculomegaly and/or hydrocephaly | + | 10/14 | 10/11 | 7/9 | 4/6 | 9/17 | 4/5 | 6/9 | 51/72 | 4/5 | 1/2 |
| Abnormality of the cerebellum | - | 0/14 | 4/11 | 1/9 | 2/6 | 5/17 | 3/5 | 3/9 | 18/72 | 2/5 | 0/2 |
| Corpus callosum abnormality | + | 4/14 | 1/11 | 5/9 | 3/6 | 8/17 | 2/5 | 7/9 | 31/72 | 3/5 | 0/2 |
| Cortical dysplasia | - | 4/14 | 1/11 | 2/9 | 2/6 | 4/17 | 1/5 | 1/9 | 15/72 | 4/5 | 0/2 |
| Seizures and/or epilepsy | + | 5/12 | 3/10 | 4/6 | 6/7 | 15/19 | 3/4 | 7/7 | 44/66 | 3/5 | 1/3 |
| Focal-onset | + | 1/5 | 2/3 | 0/4 | 4/6 | 5/15 | 2/3 | 5/7 | 20/44 | 3/3 | 0/1 |
| Generalized-onset | - | 3/5 | 1/3 | 0/4 | 5/6 | 4/15 | 2/3 | 1/7 | 16/44 | 0/3 | 0/1 |
| Hypotonia | + | 7/10 | 7/9 | 6/7 | 6/6 | 15/16 | 3/4 | 5/7 | 50/60 | 1/1 | 2/5 |
| Hypertonia | n.a. | 0/3 | 1/5 | 0/1 | 0/1 | 2/11 | 2/4 | 2/4 | 7/29 | 0/0 | 0/3 |
| Ataxia | n.a. | 2/4 | 1/5 | 1/1 | 1/3 | 2/10 | 2/3 | 2/3 | 11/29 | 1/1 | 0/3 |
| Torticollis | n.a. | 2/3 | 0/5 | 0/1 | 1/3 | 3/12 | 1/4 | 1/3 | 8/31 | 0/0 | 0/3 |
| Balance problems | n.a. | 3/4 | 2/6 | 2/2 | 2/3 | 11/13 | 3/4 | 2/3 | 25/35 | 0/0 | 1/3 |
| Developmental delay (Table 3) | + | 14/16 | 9/11 | 5/6 | 6/6 | 15/15 | 4/4 | 6/6 | 60/65 | 4/4 | 2/5 |
| Sleeping problems | n.a. | 1/3 | 4/7 | 0/1 | 3/4 | 7/14 | 2/4 | 1/3 | 18/36 | 0/0 | 2/4 |
| Insomnia | n.a. | 1/1 | 4/4 | 0/0 | 1/3 | 4/7 | 2/2 | 0/1 | 12/18 | 0/0 | 2/2 |
| Social behaviour | n.a. | 2/6 | 4/6 | 1/3 | 3/5 | 10/15 | 2/4 | 3/3 | 25/42 | 0/0 | 3/3 |
| Helpful behaviour | n.a. | 1/6 | 2/6 | 1/3 | 2/5 | 5/15 | 1/4 | 0/3 | 12/42 | 0/0 | 1/3 |
| Easily upset | n.a. | 3/6 | 3/6 | 1/3 | 2/5 | 4/15 | 2/4 | 2/3 | 17/42 | 0/0 | 1/3 |
| Hyperactivity | n.a. | 3/6 | 1/6 | 1/3 | 1/5 | 6/15 | 1/4 | 2/3 | 15/42 | 0/0 | 2/3 |

\*For explanation see Figure 1.
CT = computed tomography, MRI = magnetic resonance imaging, n.a. = not available
Clinical characteristics were selected based on prevalence and clinical significance. More details are presented in Supplementary Table S2

**Table 3.** Development for different subgroups of terminal 6q deletions

|  | T-PSMB1 | T-DLL1 | T-AFDN | T-PDE10A | T-QKI | T-PRKN | T-MAP3K4 | T-R | Total<br>(terminal) | I-6q27 | I-6q26 |
| --- | --- | --- | --- | --- | --- | --- | --- | --- | --- | --- | --- |
| Normal | 0 | 2 | 2 | 1 | 0 | 0 | 0 | 0 | 5 | 0 | **3** |
| Borderline | 0 | 1 | 1 | 0 | 1 | 2 | 0 | 0 | 5 | 0 | 1 |
| Mild delay | 0 | 2 | **3** | **2** | 1 | **6** | **1** | **2** | **17** | **4** | 1 |
| Moderate delay | 0 | **6** | 1 | 1 | **3** | 2 | **1** | **2** | 16 | 0 | 0 |
| Severe delay | 0 | 0 | 0 | 0 | 1 | 1 | 0 | 0 | 2 | 0 | 0 |
| Delayed but not specified | 1 | 5 | 4 | 2 | 0 | 4 | 2 | 2 | 20 | 0 | 0 |
| Unknown due to young age | 0 | 6 | 5 | 4 | 1 | 5 | 2 | 4 | 27 | 1 | 1 |

Not specified = developmental delay is reported, but lacking sufficient information to classify reliably. See also Figure S1
Development categorised as normal (IQ >85), borderline (IQ 70-85), mild (IQ 50-70), moderate (IQ 30-50) or severe (IQ <30) delay. The category with most individuals is highlighted in bold

**Figure 2.**
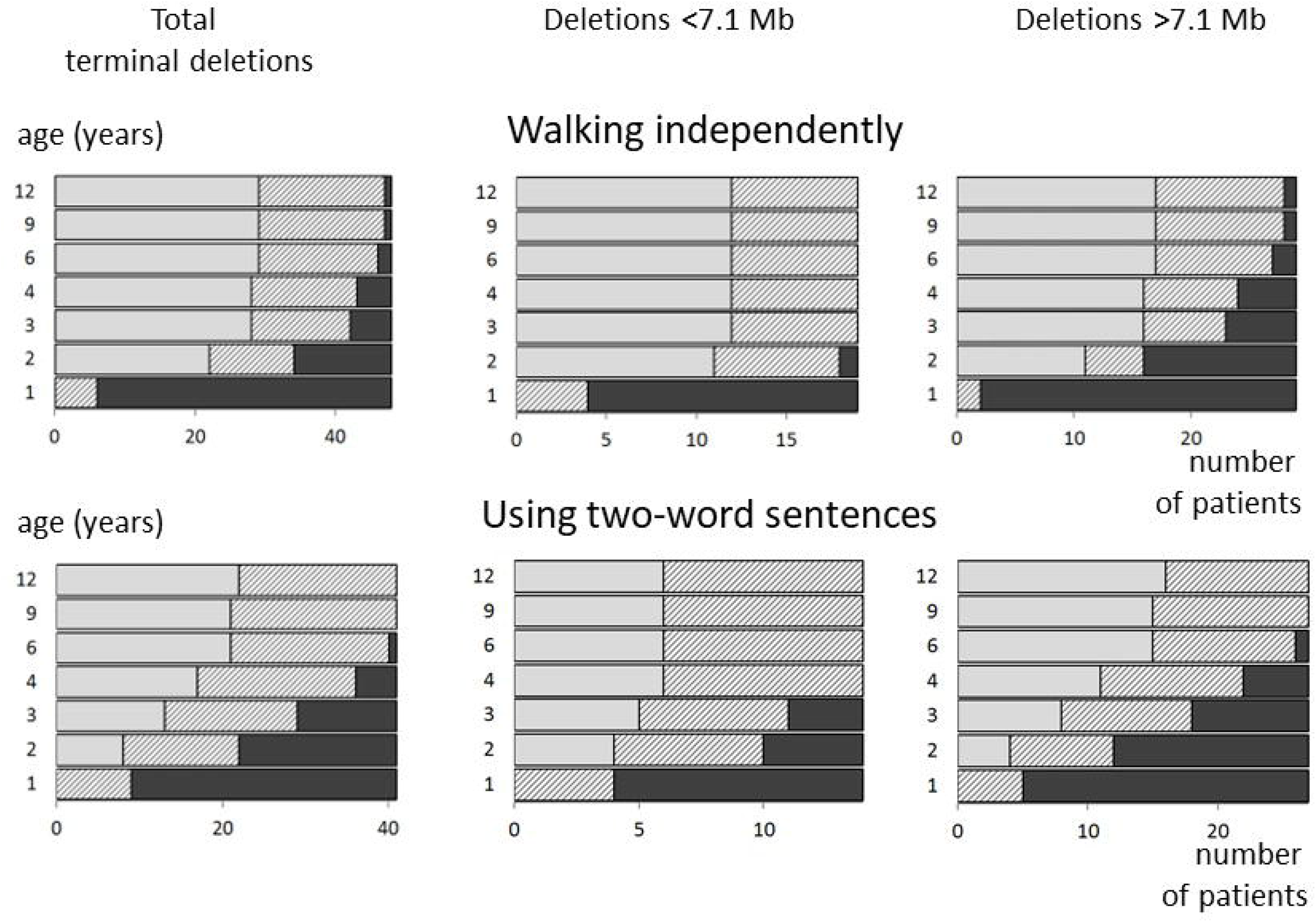
Age of achievement for milestones ‘walking independently’ and ‘using two-word sentences’ in participants who were at least 12 months of age. Deletions smaller than 7.1 Mb include subgroups T-PSMB1, T-DLL1, T-AFDN and T-PDE10A. Deletions larger than 7.1 Mb include subgroups T-QKI, T-PRKN, T-MAP3K4 and T-R. Light grey bars indicate the number of children (x axis) that have reached the milestones ‘walking independently’ (upper panel) and ‘using two-word sentences’ (lower panel) before the given age (y axis, years). The dark grey bars are the children who were not able to perform the milestone at that age. The hatched bars are the children who were not able to perform the milestone, but who have not yet reached the age on the y axis. For example, at age 12 years, 29– 47 (60–98%) participants are able to walk (figure and description adapted from Engwerda, et al. 2018^4^).

### The terminal 6q deletion phenotype

The pregnancy was characterised by intrauterine growth restriction in 30% of individuals and reduced foetal movements in 26% of individuals. Prolonged neonatal jaundice was reported in 9/30 new-borns with a terminal deletion including *PDE10A* (larger than 5 Mb).

Many individuals had a small head size (<p10) (60%) and/or plagiocephaly (47%). Some individuals had a short body stature (<p10) (30%). Dysmorphisms included hypertelorism (51%), dysplastic ears (51%) and dental abnormalities (38%). A cleft lip and/or palate was seen in three individuals with larger deletions, see Table S2. One individual (T-R) also had a choanal stenosis.

Decreased visual acuity (67%) was mostly mild (16/30). Abnormal eye movements (56%) included strabismus (24/30) and nystagmus (11/30). Aplasia/hypoplasia of the optic nerve was seen in 4/26 individuals with a deletion including at least *PRKN* (larger than 7.9 Mb). Microphthalmos (2/4) and coloboma (1/4) were only seen with the largest deletions, subgroup T-R (larger than 10.5 Mb).

Feeding problems (72%) were common, with oral aversion (61%), chewing difficulties (43%), dysphagia (39%) and gastroesophageal reflux (26%) most often reported. Seven individuals needed a gastrostomy, and all seven had a deletion including *PRKN*. Bowel problems (43%), often constipation (32%), were also reported, but never for the smallest deletions, subgroup T-DLL1 (smaller than 2.7 Mb). Five out of 24 individuals with a deletion including the gene *QKI* (larger than 7.1 Mb) had an abnormality of the anus, either anal atresia (n=3) or an ectopic anus (n=1).

Recurrent infections (42%) were often reported, including of the upper respiratory tract (13/13). Respiratory problems were reported in 26% of the individuals. Sleep apnoea (n=3) was only present in deletions including at least *QKI*. Congenital heart defects (CHDs) (12/42), kidney problems (7/37) and abnormalities of the female genitalia (6/27) were only reported in individuals with a deletion including *AFDN* (larger than 2.7 Mb).

A sacral dimple was reported in 23/31 individuals with a deletion including *AFDN*. Spina bifida was seen in 5/21 individuals with a deletion including *PRKN* (larger than 7.9 Mb). Scoliosis (6/39) and abnormal vertebrae (8/38, including hemivertebrae in n=4) were seen throughout the whole group. Joint hypermobility was present in 64% of all individuals, and hip dysplasia was reported in 6/15 individuals with a *PDE10A* deletion. In individuals with a deletion including *AFDN*, a positional foot deformity (8/14) and pes planus (5/14) were reported.

Almost all individuals had brain abnormalities on MRI or CT (91%). Those most often reported were ventriculomegaly/hydrocephaly (51/72), corpus callosum abnormalities (31/72), abnormalities of the cerebellum (18/72) and cortical dysplasia (15/72). In literature, other brain abnormalities were reported that were not included in the Chromosome 6 Questionnaire, these included colpocephaly (n=8), periventricular nodular heterotopia (PNH) (n=6), hypoplasia of the hippocampi (n=5) and a large cisterna magna (n=5), and all were seen throughout all subgroups^7,9,20,26,28^. Seizures (67%) (generalised (16/44) and focal onset (20/44)) were often reported, and epilepsy was formally diagnosed in 83% of individuals with seizures. Hypotonia (83%) was very common. Hypertonia was reported in 7/29 individuals, and most of these individuals had a deletion including at least *PRKN*. 3/17 individuals with a *PRKN* deletion were reported with spasticity. Balance problems (71%) were often seen. Torticollis (26%), ataxia (38%) and spinal cord abnormalities (39%) were reported throughout the whole group, as well as abnormal pain sensation (30%) and sensory integration disorder (30%).

Behaviour was most often described as being social (60%), helpful (29%), easily upset (40%) and hyperactive (36%). Seven out of 22 individuals with a *PRKN* deletion showed self-harming behaviour. Half of the individuals had sleeping problems (50%), most frequently insomnia (67%).

Almost all individuals had developmental delay (92%), see Table 3 and Figure S1. Five individuals had no developmental delay, and their ages ranged from 5–49 years and their deletion sizes from 2–5.5 Mb, see Figure S1^16,20^. The level of developmental delay varied considerably within all the subgroups, but was mostly mild to moderate. Individuals without a deletion of *QKI* (deletion smaller than 7.1 Mb) had normal development to moderate delay, and individuals with a deletion including *QKI* (larger than 7.1 Mb) had borderline to severe developmental delay.

Ages of achievement for certain milestones are summarised in Table 4 and visualised in Figure 2. All children with a deletion not including *QKI* (smaller than 7.1 Mb) were able to perform the milestone ‘walking independently’ at age 3 years and the milestone ‘using two-word sentences’ at age 4 years. For individuals with larger deletions including *QKI* (larger than 7.1 Mb), some needed more time to achieve these milestones and one individual was not yet able to walk independently at age 12 years. The milestones ‘roll over’, ‘sit up unaided’ and ‘pull up in a standing position’ were achieved significantly earlier by children with deletions that did not include *QKI*. Five out of 22 children reached the milestone ‘fully toilet trained during the day’ at age 4 years.

**Table 4.** Age of achievement for milestones

| Milestones | Total terminal deletions |  | Deletions <7.1 Mb |  | Deletions >7.1 Mb |  |
| --- | --- | --- | --- | --- | --- | --- |
|  | Milestone achieved | Not able to perform milestone | Milestone achieved | Not able to perform milestone | Milestone achieved | Not able to perform milestone |
| Roll over (weeks) | n=30<br>9 (2–28) | n=2<br>27.5 (25–30) | n=9<br>8 (4–10)* | n=1<br>25 | n=21<br>10 (2–28)* | n=1<br>30 |
| Sit up unaided (months) | n=29<br>13 (7–40) | n=5<br>10 (6–28) | n=11<br>11 (7–18)* | n=2<br>8 (6–10) | n=18<br>15 (9–40)* | n=3<br>15 (7–28) |
| Pull up in a standing position (months) | n=24<br>16.5 (9–72) | n=10<br>15 (6–57) | n=6<br>12.5 (9–18)* | n=4<br>11 (6–16) | n=18<br>18 (11–72)* | n=6<br>17 (7–57) |
| Walking unsupported (months) | n=28<br>22.5 (14–57) | n=17<br>19 (6–393) | n=12<br>18 (14–30) | n=5<br>12 (6–16) | n=16<br>23.5 (15–57) | n=12<br>30.5 (7–393) |
| Speak first words (months) | n=25<br>14 (9–60) | n=11<br>15 (6–86) | n=6<br>15.5 (10–30) | n=6<br>14 (6–44) | n=19<br>13 (9–60) | n=5<br>15 (7–86) |
| Speak two-word sentences (months) | n=19<br>30 (18–72) | n=16<br>19.5 (6–86) | n=6<br>24 (20–45) | n=6<br>14 (6–44) | n=13<br>36 (18–72) | n=10<br>24 (7–86) |
| Fully toilet trained during the day (years) | n=10<br>4.5 (3–10) | n=27<br>3.5 (0.5–32) | n=3<br>4 (3–4) | n=7<br>3.5 (0.5–8) | n=7<br>5 (4–10) | n=20<br>4 (0.5–32) |

For each milestone the number of individuals who achieved the milestone and the median and range of their ages of achievement are given. The number of individuals who were not able to perform the milestone is also given and the median and range of the age at which they could not yet perform the milestone. Deletions smaller than 7.1 Mb include subgroups T-PSMB1, T-DLL1, T-AFDN and T-PDE10A, deletions larger than 7.1 Mb include subgroups T-QKI, T-PRKN, T-MAP3K4 and T-R.
\* Significant ( $p < 0.05$ ) difference between the age of achievement for smaller (<7.1 Mb) and larger (>7.1 Mb) terminal deletions.

Two out of seven index patients with an inherited deletion (subgroups T-DLL1 and T-AFDN), inherited this from an independently functioning parent (1 maternal, 1 paternal)^20^.

Nineteen out of 93 individuals had reached adulthood, and their median age was 32 years (range 18–57). Detailed information on adult functioning could only be collected for two adults from the parent cohort. One individual needed help with all tasks and was not able to take care of herself (T-PRKN). The other adult (T-PRKN) was able to perform simple tasks (for example handling a phone and performing some daily housework) but needed assistance with most tasks.

Some clinical characteristics were reported in literature that were not part of the Chromosome 6 Questionnaire. In addition to the brain abnormalities reported above, micrognathia and high arched palate were reported in 13 and 14 individuals in literature, respectively^1,7–10,13,19,23,26,28^. These characteristics could also not be related to deletion size.

### Interstitial 6q26 and 6q27 deletion phenotype

All five interstitial 6q27 deletion individuals were derived from literature, and the only deleted HI- gene they had in common was *DLL1* (Figure 1). Their phenotype was mostly characterised by brain abnormalities (ventriculomegaly/ hydrocephaly, corpus callosum abnormality and cortical dysplasia), epilepsy and mild developmental delay. Other characteristics reported were small head size, strabismus, horseshoe kidney, joint hypermobility, hypotonia, ataxia and autistic behaviour.

There were three interstitial 6q26 deletion individuals in the parent cohort and another three in the literature cohort. *PRKN* was involved in all deletions, and the largest deletion also encompassed *QKI* and *PDE10A* (Figure 1). Individuals with 6q26 interstitial deletions were mostly characterised by a normal head size and normal body stature, brain abnormalities (delayed myelination, ventriculomegaly/hydrocephaly), vision problems, feeding problems, an abnormal curvature of the spine (scoliosis/kyphosis), hypotonia and insomnia. Other characteristics reported were atrial septal defect (ASD), hydronephrosis, constipation, spina bifida, epilepsy and sleep apnoea. Their behaviour was mostly characterised by social behaviour, hyperactivity, attention deficit disorder and autistic behaviour/disorder. One individual presented self-harming behaviour. Three individuals had no developmental delay, one had borderline delay and one had mild delay (Figure S1 and Table 3).

### Ring 6 phenotype

For two individuals, their terminal deletion was known to be the result of a ring chromosome 6^23,32^. The ring also included a small terminal 6p deletion without phenotypic consequences^38^. There were no differences in the phenotype between the individuals with a ring chromosome and individuals with a comparable simple deletion.

### Effect of the common fragile site *FRA6E*

None of the 93 terminal deletions had a start breakpoint within the fragile site *FRA6E* (Figure 1).

## Discussion

Here we have reported our findings on rare chromosome 6q deletions in the region 6q26q27, which are mainly terminal 6q deletions. Below, we summarise the most clinically relevant characteristics and provide advice about additional medical examinations, follow-up and surveillance. We also briefly discuss the genes involved in the deletions and, based on what is known about these genes, how they could be linked to the observed clinical characteristics. We found that there is only a minor effect of deletion size on the clinical phenotype, resulting in a common (but variable) phenotype seen in all terminal 6q deletion individuals. This commonality could be explained by the prominent role of the distally located gene *DLL1*, with only individuals with deletions larger than 7.1 Mb clearly showing additional or more severe clinical characteristics.

In our cohorts there is a clear common terminal 6q deletion phenotype characterised by a small head size, dysplastic outer ears, hypertelorism, vision problems, abnormal eye movements, dental abnormalities, feeding problems, recurrent infections, respiratory problems, spinal cord abnormalities, abnormal vertebrae, scoliosis, joint hypermobility, brain abnormalities (ventriculomegaly/hydrocephaly, corpus callosum abnormality and cortical dysplasia), seizures, hypotonia, ataxia, torticollis, balance problems, developmental delay, sleeping problems and hyperactivity. Since these characteristics are common in the affected individuals, it is most likely that the phenotype is caused by haploinsufficiency of the most distally located genes. Nonetheless, the phenotype is very variable, and not all characteristics are seen in all individuals.

The three most distally located HI-genes that could contribute to this common 6q terminal deletion phenotype are *TBP* (Tata Box-Binding Protein, MIM*600075), *PSMB1* (Proteasome Subunit Beta-Type 1, MIM*602017) and *DLL1. TBP* is known to cause late-onset neurological disorders such as spinocerebellar ataxia (MIM#607136) and Parkinson’s disease (MIM#168600) through expansion of a CAG repeat^39^, but it is unclear whether loss of function of one allele has a phenotypic affect. Mice haploinsufficient for *Tbp* did not show any abnormalities^40^, and no pathogenic loss-of-function variants have been reported in humans. For the gene *PSMB1*, no pathogenic heterozygous variants have been reported. Recently, a presumed pathogenic homozygous missense variant was reported resulting in microcephaly, developmental delay and short stature in two sisters, aged 22 and 35 years^41^. These clinical characteristics are present in our cohort but also appear in individuals with an interstitial 6q27 deletion that did not involve *PSMB1*^*7,29*^. Furthermore, there are multiple individuals with a deletion including both *TBP* and *PSMB1* in the database of genomic variance (DGV)^42^. The presence of these deletions in people with a normal or unrelated phenotype makes it unlikely that *TBP* or *PSMB1* play a major role in the terminal 6q deletion phenotype.

### DLL1

The third distally located HI-gene of interest for the common 6q terminal deletion phenotype is *DLL1*. Recently, Fischer-Zirnsak et al. (2019) described 14 patients with pathogenic heterozygous variants of *DLL1* and one patient with a deletion including *DLL1*. These patients presented with hypotonia, scoliosis and a neurodevelopmental phenotype including variable brain abnormalities (ventriculomegaly/hydrocephaly, corpus callosum abnormality and cortical dysplasia), seizures and autism spectrum disorder^29^. This phenotype was registered in OMIM as neurodevelopmental disorder with nonspecific brain abnormalities and with or without seizures (NEDBAS, OMIM#618709). Additionally, the following clinical characteristics were reported in at least one patient with a heterozygous *DLL1* variant: PNH, large cisterna magna, strabismus, feeding problems, sleep apnoea, recurrent infections, hemivertebrae, sacral dimple, joint hypermobility, ataxia and hyperactivity^29^. As all these clinical characteristics are also seen in our terminal 6q deletion cohorts, it is very likely that haploinsufficiency of *DLL1* makes an important contribution to the common terminal 6q deletion phenotype. Lesieur-Sebellin et al. characterised the features detected by prenatal ultrasound in 22 foetuses with terminal 6q deletions and pointed out the gene *DLL1* as the major contributor to the detected phenotype^43^.

In 2005, Eash et al. reported a patient (Eash_1) with the smallest terminal deletion seen thus far, 390kb, which only included the HI-genes *TBP* and *PSMB1*^*1*^. This patient’s phenotype was comparable to the common terminal 6q deletion phenotype we describe and included microcephaly, brain abnormalities (corpus callosum abnormality, hydrocephaly), seizures, vertebral abnormalities, hypotonia and developmental delay (Tables 2, 3 and S2; subgroup T-PSMB1). Most other characteristics of the common terminal 6q deletion phenotype were not reported as being present or absent in the Eash et al. case. The breakpoints of this deletion where defined by BAC and PAC FISH clones at approximately 500 kb intervals^1^. If the possible maximum size of the deletion is taken into consideration (Figure 1), the *DLL1* gene could actually be part of the deletion. We tried contacting the authors about additional genetic studies performed for this individual, but without success. Considering the overlapping phenotype and the ambiguities in breakpoint definition, we expect that *DLL1* is also part of, or influenced by, the deletion in this case.

*DLL1* codes for a ligand of the Notch receptor. The Notch signalling developmental pathway is involved in cell-to-cell communication and cell patterning and differentiation. Notch signalling plays a role in the development of multiple organs and tissues, including the somite-derived organs, nervous system, heart, vasculature, haematopoietic system, cochlea and pancreas^44^. In our cohorts, we did not clearly see any abnormalities for the four latter organ systems, but the nervous system, somite-derived organs and heart were affected.

In mice, delayed expression of *Dll1* leads to premature cell differentiation, resulting in a smaller brain and fused vertebrae and ribs^45^. This is in line with the variable brain abnormalities, microcephaly and abnormalities of the vertebrae seen in our cohorts and in the heterozygous *DLL1* variant patients reported by Fischer-Zirnsak et al.^29^.

CHDs were not reported in the patients by Fischer-Zirnsak et al.^29^ and also not seen in our T- DLL1 subgroup, so the effect on heart development may not be fully penetrant. We did find CHDs in larger deletions that include *DLL1*. In these patients, *DLL1* seems the most likely candidate gene to cause CHDs given its role in the Notch pathway and reported pathogenic variants in *NOTCH1* in patients with a CHD^46^. During heart development, Notch signalling plays a crucial role in the formation of the membranous walls of the atrial and ventricular chambers and of the outflow tract^47^. Interruption in Notch signalling could explain the CHDs in our cohorts: ASDs, VSDs and a coarctation of the aorta in one individual. Bu et al. reported a patient with an ASD and persistent left superior vena cava with a heterozygous *DLL1* variant that was classified as likely pathogenic. This *DLL1* variant patient also had a cleft palate, but no further phenotype information was given^48^. A cleft palate was also seen in two of our patients, although these two had the largest terminal deletions of our cohort.

### Other genes

Besides *DLL1*, other genes were also previously proposed to play a role in the terminal 6q deletion phenotype, especially in larger terminal deletions that display additional clinical characteristics. Below, we briefly summarise these in the context of our findings.

Several genes have been linked to brain abnormalities. *ERMARD* (Endoplasmic Reticulum Membrane-Associated RNA Degradation Protein, MIM*615532 (also known as C6orf70)) might be involved in PNH, since Conti et al. described nine patients with a deletion including *ERMARD* and one patient with a heterozygous missense variant and PNH^7^. Unfortunately, we do not have information on the prevalence of PNH for our parent cohort. One patient in our literature cohort did present with PNH, but her deletion did not include *ERMARD*^*28*^. *ERMARD* is also not a predicted HI-gene (%HI: 84.86, pLI: 0.00). Based on this information and the fact that the deletion patients presenting with PNH of Conti et al. all had a deletion that also included *DLL1*, it remains unclear whether haploinsufficiency of *DLL1* or *ERMARD*, or both, can cause PNH.

Peddibhotla et al. suggested two other genes that may be involved in structural brain abnormalities: *THBS2* (Thrombospondin II, MIM*188061) and *PHF10* (Phd Finger Protein 10, MIM*613069). These genes were deleted in all seven of their terminal 6q deletion patients^9^. However, both genes are not predicted HI-genes, and no pathogenic variants causing structural brain abnormalities in humans have been described thus far.

Lastly, *QKI* has been linked to brain abnormalities because it plays a role in myelination by regulating several myelin-specific genes^49^. Five individuals in our terminal 6q deletion cohort presented with delayed myelination, and all have a *QKI* deletion (Figure S2). One individual with an interstitial 6q26 deletion also presents with delayed myelination, but *QKI* is not part of her deletion, and 32 out of 37 patients with a deletion of *QKI* did not have delayed myelination. Likewise, Backx et al. reported a woman with a reciprocal balanced translocation t(5;6)(q23.1;q26) disrupting the *QKI* gene, resulting in 50% reduced QKI expression, who did not present with myelination problems^50^. Thus, although it is likely that *QKI* plays a role in myelination, there seems to be incomplete penetrance of this clinical feature.

Vertebral abnormalities are part of the common terminal 6q deletion phenotype, and *TBXT* (T-Box Transcription Factor T, MIM*601397) is suggested to play a role in the aetiology of hemivertebrae^18^. An identical missense variant in this gene was identified in three unrelated patients with congenital vertebral malformations. This variant was proposed to increase the risk of congenital vertebral malformations, but not sufficiently on its own^18,51^. Nevertheless, *TBXT* was not deleted in all patients with hemivertebrae in our cohort. We therefore think this phenotype is more likely linked to *DLL1*.

Dental problems, including abnormal morphology and reduced number of teeth, are also part of the common terminal 6q deletion phenotype. The gene *SMOC2* is related to dental problems in carriers of pathogenic homozygous variants, including oligodontia, microdontia and abnormally shaped teeth^52–54^. However, no pathogenic heterozygous *SMOC2* variants have been identified thus far, and *SMOC2* was not deleted in all individuals with dental problems in our cohort (Figure S3).

CHDs were seen in 12 patients with terminal deletions including at least *AFDN* (deletions larger than 2.7 Mb). Next to *DLL1*, these deletions also included the gene *THBS2*. In two large CHD cohort studies, two variants of unknown significance in *THBS2* were found. One patient presented with a tetralogy of Fallot^55^. The other patient presented with subaortic stenosis, bicuspid aortic valve, mitral valve stenosis and regurgitation and a coarctation of the aorta^56^. In contrast, our CHD patients mainly presented with septal defects. Since there is no further proof for the role of *THBS2* in CHD, and all terminal 6q deletions also include the more likely candidate gene *DLL1*, we regard the contribution of *THBS2* to CHD in the 6q deletion phenotype as less likely.

Recent work showed that *QKI* also plays a role in cardiovascular development and function in mice and might be involved in cardiomyopathies and cardiovascular disease in humans^57^. One (1/42) of our patients (aged <5 years) with a deletion including *QKI* was reported to have hypertrophic cardiomyopathy. Further research is needed to investigate whether there is a relation between cardiomyopathies and a deletion of *QKI* and thus whether individuals with a *QKI* deletion need to be screened for cardiomyopathy.

Almost all the individuals in our cohort with a deletion including *DLL1* had developmental delay. Besides *DLL1, QKI* seems to mark a tipping point in the extent of developmental delay. Normal development was seen in a couple of individuals without a deletion of *QKI*, whereas severe developmental delay was only seen in individuals with a deletion including *QKI* (Figure S1, Table 3). The woman with a reciprocal balanced translocation t(5;6)(q23.1;q26) disrupting the *QKI* gene reported by Backx et al. also presented with borderline developmental delay^50^. *QKI* probably has an additive effect on the level of developmental delay next to the deletion of *DLL1*, which on its own can lead to moderate developmental delay in small (500 kb) deletions^28^.

A range of behavioural problems was seen throughout the whole group of terminal deletions, with information available for all 34 individuals from the parent cohort but only 8 of 51 literature cases, foetuses excluded (Table S2). Self-harming behaviour was seen significantly (Fisher’s Exact Test p=0.03) more often in the larger terminal deletions. Fischer et al. reported autism spectrum disorder as part of the syndrome linked to *DLL1* haploinsufficiency. In their cohort, 5 out of their 13 *DLL1* variant patients and their one *DLL1* deletion patient had autism spectrum disorder^29^. In our cohorts, however, autistic behaviour was present in only 4/33 individuals with a terminal deletion including *DLL1*. However, autistic behaviour was also seen in 3/5 individuals with an interstitial 6q26 deletion that did not include *DLL1*, suggesting that *DLL1* haploinsufficiency is not the only cause for autistic behaviour.

The individuals with terminal 6q deletions included in our cohort were grouped based on the number of predicted HI-genes involved in their deletion. However, only two HI-genes, *DLL1* and *QKI*, could be linked to some of the observed clinical characteristics. This is reflected in the phenotypic differences between the patients with deletions smaller and larger than 7.1 Mb.

### FRA6E

It has been suggested that the *FRA6E* fragile site is the cause of the breakpoints in terminal 6q deletions^3,12,13^. *FRA6E* is a common fragile site located at 161.71–161.91 Mb in 6q26 (Figure 1).

Common fragile sites are associated with hotspots for chromosome aberration breakpoints^58^ due to an impaired replication process at the fragile site. Nevertheless, Palumbo et al. showed that the replication process at *FRA6E* is not impaired^59^, and we also do not see a clustering of breakpoints within or near the fragile site in our data.

### Recommendations for clinical follow-up and surveillance

The common terminal 6q deletion phenotype is highly variable, and not all clinical characteristics are present in each individual. Therefore clinical follow-up and surveillance should be focused on the congenital anomalies present and the problems experienced. This also means that the phenotype should be fully assessed upon diagnosis to establish which of the comorbidities known for this chromosomal aberration are present in the patient. In Table 5, we present our recommendations for investigations for terminal 6q deletions <7.1 Mb and for deletions >7.1 Mb including the gene *QKI*. We recommend that all individuals should be offered a neurological investigation (including an MRI upon indication), a cardiac ultrasound (at least once), an investigation by an ophthalmologist and an annual vision assessment at younger ages and upon indication at older ages. As seizures are seen in a large proportion of the individuals, the threshold should be low for consulting a (paediatric) neurologist and for performing an EEG. Individuals with hypermobility can benefit from physiotherapy and medical aids. For individuals with deletions >7.1 Mb, clinicians should also be aware of the occurrence of cleft palate, anal atresia and sleep apnoea, which are reported in some individuals and can be treated. Some individuals with deletions >7.1 Mb have spina bifida, although vertebral abnormalities that could cause scoliosis are seen for all deletion sizes. Performing an X-ray of the vertebral column can help identify these abnormalities. Besides this specific advice for individuals with a terminal 6q deletion, appropriate support for feeding, behavioural and sleeping problems should also be in place.

**Table 5.**
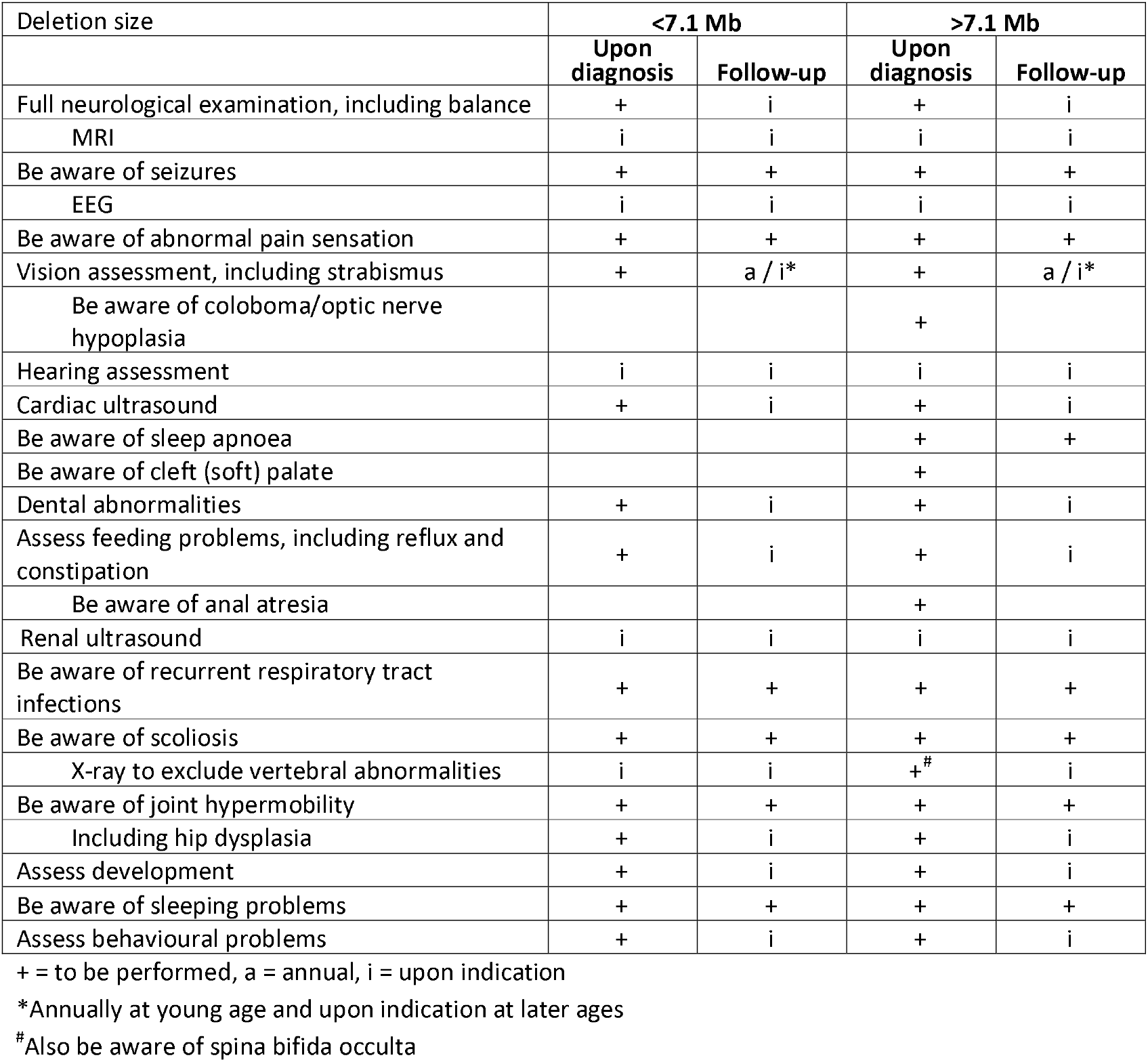
Clinical recommendations for terminal 6q deletions

| Deletion size | <7.1 Mb |  | >7.1 Mb |  |
| --- | --- | --- | --- | --- |
|  | Upon diagnosis | Follow-up | Upon diagnosis | Follow-up |
| Full neurological examination, including balance | + | i | + | i |
| MRI | i | i | i | i |
| Be aware of seizures | + | + | + | + |
| EEG | i | i | i | i |
| Be aware of abnormal pain sensation | + | + | + | + |
| Vision assessment, including strabismus | + | a / i* | + | a / i* |
| Be aware of coloboma/optic nerve hypoplasia |  |  | + |  |
| Hearing assessment | i | i | i | i |
| Cardiac ultrasound | + | i | + | i |
| Be aware of sleep apnoea |  |  | + | + |
| Be aware of cleft (soft) palate |  |  | + |  |
| Dental abnormalities | + | i | + | i |
| Assess feeding problems, including reflux and constipation | + | i | + | i |
| Be aware of anal atresia |  |  | + |  |
| Renal ultrasound | i | i | i | i |
| Be aware of recurrent respiratory tract infections | + | + | + | + |
| Be aware of scoliosis | + | + | + | + |
| X-ray to exclude vertebral abnormalities | i | i | +<sup>#</sup> | i |
| Be aware of joint hypermobility | + | + | + | + |
| Including hip dysplasia | + | i | + | i |
| Assess development | + | i | + | i |
| Be aware of sleeping problems | + | + | + | + |
| Assess behavioural problems | + | i | + | i |

+ = to be performed, a = annual, i = upon indication
\*Annually at young age and upon indication at later ages
<sup>#</sup>Also be aware of spina bifida occulta

Development was delayed in most individuals, and those with deletions >7.1 Mb needed more time to achieve developmental milestones than those with deletions <7.1 Mb. Nonetheless, all but one individual did achieve the milestones ‘walking independently’ and ‘using two-word sentences’ (Table 4 and Figure 2). As in all neurodevelopmental disorders, the definite aim should be to optimise the developmental abilities and quality of life of the individual with a chromosomal aberration and their family. Having more detailed information available on what to expect is an important step in this process.

### Limitations of the study and suggestions for future research

Recruiting parents via social media and collecting phenotypes directly from the parents via the online Chromosome 6 Questionnaire resulted in an extensive dataset for the parent cohort. However, not all clinical characteristics were addressed in literature, resulting in missing data. For example, it was only known for one out of ten (10%) individuals in the T-PDE10A subgroup if they had sleeping problems, while this was known for 14 out of 20 (70%) in the T-PRKN subgroup. The main difference between these subgroups was the ratio of parent cohort versus literature cohort cases, which was 1:9 for T-PDE10a and 15:5 for T-PRKN. In another study, we investigated the availability of data on specific phenotype information in literature case reports compared to data collected directly from parents in the terminal 6q deletion cohort presented here. We show that we collected significantly more data from parents, for almost all phenotypic features, in comparison to the literature^60^.

A risk of not reporting data on absent phenotype features is that incorrect conclusions can be drawn. For example, balance problems were often reported in the whole terminal 6q deletion cohort (25/35, 71%), but vestibular and/or cerebellar dysfunction was only reported as a cause of these balance problems in individuals with a deletion including *PRKN* (larger than 7.9 Mb) (5/16, 31%). It remains unknown whether vestibular and/or cerebellar dysfunctions only cause balance problems in patients with larger deletions, or if the causes of balance problems in those with smaller deletions were simply not investigated or reported.

Our phenotype data was collected directly from parents, which might raise questions on the quality of the data. However, in our data consistency study we show that phenotype data collected directly from parents is highly consistent with data extracted from the medical files on the same individual^60^.

Another topic for which information is still very limited is the natural history of disease and adulthood. For two individuals with a terminal 6q deletion smaller than 2.7 Mb, it is known that they did not have developmental delay and could live independently. For another two individuals with a deletion larger than 7.9 Mb (including *PRKN*), it is known that they could not (fully) take care of themselves and could not live independently. However, for most individuals who have reached adulthood, information on their level of performance is very limited and it was unclear whether they could live independently. Follow-up on adults with terminal 6q deletions is needed to give insight into adult functioning and development of new clinical features at older ages.

Our recommendations for investigations now focus on two groups – deletions <7.1 Mb and deletions >7.1 Mb including the gene *QKI* – since this was a clear tipping point in the reported phenotypes. However, we cannot be absolutely sure that these clinical characteristics are only seen in the larger deletions and will never be reported in individuals with smaller deletions. Therefore we have tried to be cautious in our recommendations for investigations. Since the phenotype can be very variable, it is important to assess each patient on an individual level. Nonetheless, the general differences we have reported can be helpful in counselling (expecting) parents. For the future, we hope to be able to give more detailed recommendations based on deletion sizes, but this is only possible if detailed information for an even larger study population is collected.

## Conclusions

Terminal 6q deletions cause a common phenotype that is broad and highly variable within individuals and within families. The main clinical characteristics are microcephaly, brain abnormalities (ventriculomegaly/hydrocephaly, corpus callosum abnormalities and cortical dysplasia), neurological problems (seizures, hypotonia and ataxia), vision problems, developmental delay, behavioural problems and subtle dysmorphic features. Cardiac, gastrointestinal, urogenital and skeletal anomalies may also occur. Most of the characteristics can be linked to the distally located gene *DLL1* and, probably as a consequence, deletion size has little effect on the phenotype. However, individuals with deletions >7.1 Mb that include *QKI* present with a more severe phenotype that includes severe developmental delay. Based on our findings, we provide recommendations for clinical follow-up and surveillance of individuals with terminal 6q deletions. To further improve these recommendations, more data needs to be collected, especially on clinical follow-up and adult functioning.

## Data Availability

All data produced in the present study are available upon reasonable request to the authors

https://decipher.sanger.ac.uk

## List of abbreviations

ASD: atrial septal defect
ADHD: attention deficit hyperactivity disorder
CHD: congenital heart defect
DGV: database of genomic variance
HI: haploinsufficiency
pLI: loss-of-function intolerance
PNH: periventricular nodular heterotopia

## Declarations

### Ethics approval and consent to participate

The accredited Medical Ethics Review Committee of the University Medical Center Groningen waived full ethical evaluation because, according to Dutch guidelines, no ethical approval is necessary if medical information that was already available is used anonymously and no extra tests have to be performed.

### Consent for publication

Consent for publication was obtained from all participants, and additional consent was obtained for the use of photographs.

### Availability of data and materials

Data collected from individuals in the parent cohort was submitted to the DECIPHER database (decipher.sanger.ac.uk), IDs 489709–489746.

### Authors’ contributions

Conceptualisation: AE, CMAR; Data curation: AE; Funding acquisition: AE, CMAR; Investigation: AE, CMAR; Methodology: AE, CMAR; Project administration: AE; Resources: AE, CMAR; Software: AE; Supervision: CMAR; Visualisation: AE; Writing – original draft: AE; Writing – review & editing: AE, WSK, NC, TD, CMAR.

## Acknowledgements

We would like to thank all the children and families for their participation. We also thank Kate Mc Intyre for editing the manuscript, Nadia Simoes de Souza for helping with data collection and first analysis, Dr. Valerio Conti for providing patient information and Morris Swertz and the members of the MOLGENIS team at the Groningen Genomic Coordination Center for their support on the online Chromosome 6 Questionnaire. Our special thanks go to Pauline Bouman, our contact for the Chromosome 6 Facebook group.

## Supplementary files

Figure S1. Developmental delay in children older than 2 years of age

Figure S2. Delayed Myelination

Figure S3. Dental problems

Table S1. HI and pLI scores

Table S2. Phenotypes detailed

