## Supplementary files for "The phenotypic spectrum of terminal 6q deletions based on a large cohort derived from social media and literature: a prominent role for *DLL1*"

**Supplementary Figures**

Figure S1. Developmental delay in children older than 2 years of age
Figure S2. Delayed Myelination
Figure S3. Dental problems

**Supplementary Table**

Table S1. HI and pLI scores

**Figure S1. Developmental delay in children older than 2 years of age**

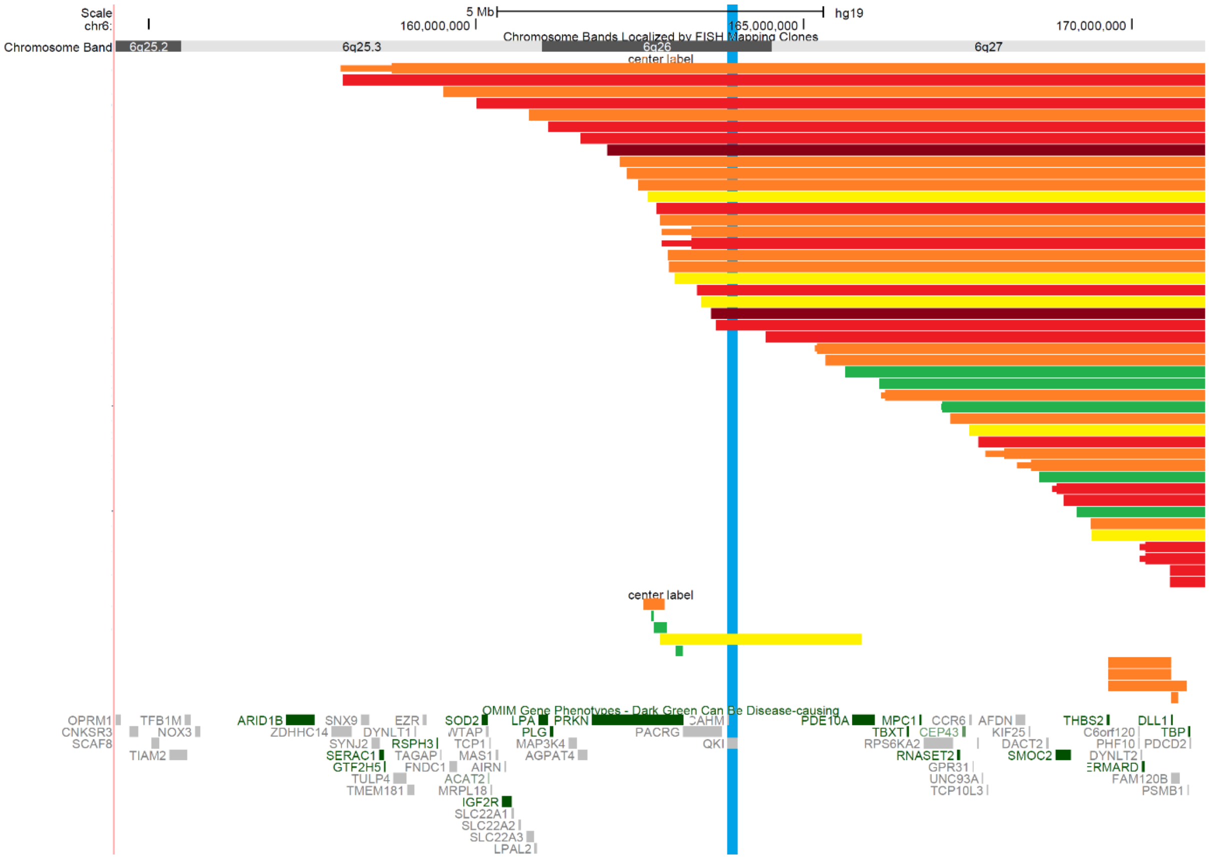


The deletion in each patient for whom development could be categorised is represented by a horizontal bar. Development is categorised as normal (IQ >85, green bar), borderline (IQ 70-85, yellow), mild (IQ 50-70, orange), moderate (IQ 30-50, red) or severe (IQ <30, dark red) delay. The gene *QKI* is represented by a vertical blue bar (see manuscript Discussion).

### Figure S2. Delayed Myelination

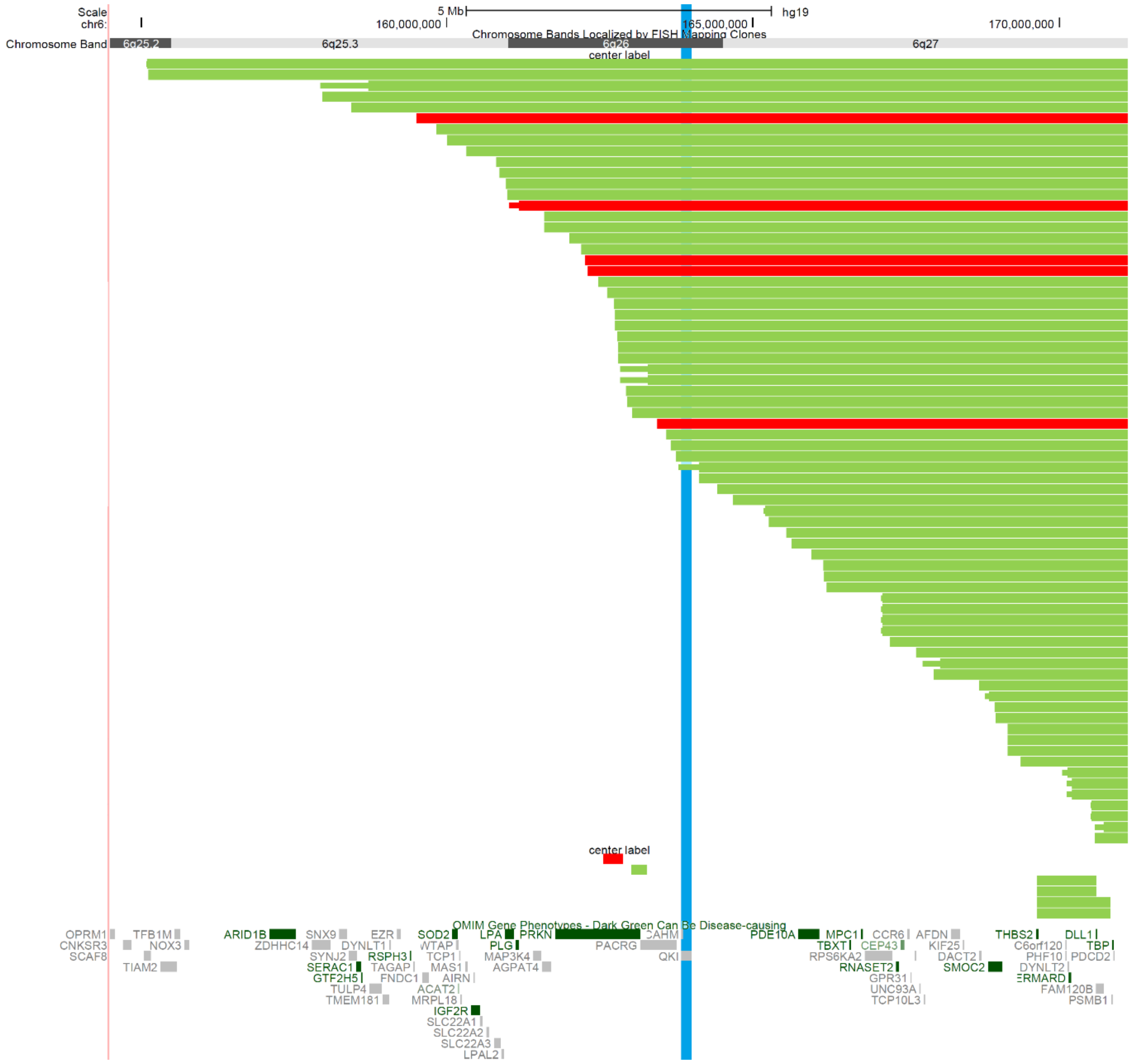


The deletion in each patient for whom delayed myelination was present (red) or absent (green) is represented by a horizontal bar. The gene *QKI* is represented by a vertical blue bar (see manuscript Discussion).

### Figure S3. Dental problems

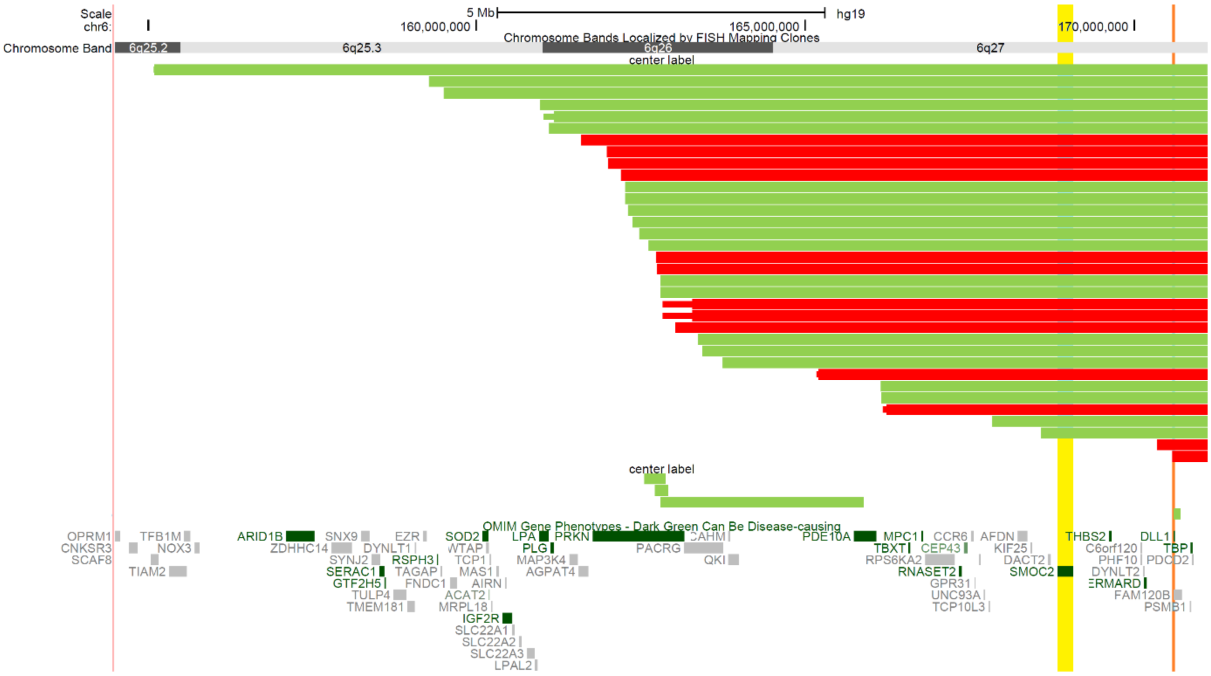


The deletion in each patient for whom dental problems were present (red) or absent (green) is represented by a horizontal bar. The gene *SMOC2* is represented by a vertical yellow bar and the gene *DLL1* is represented by a vertical orange bar. *SMOC2* is related to dental abnormalities in pathogenic homozygous variants (see manuscript Results and Discussion).

**Table S1. HI and pLI scores**

| Location | Gene | MIM* | % HI | pLI |
|---|---|---|---|---|
| 6q27 | *PDCD2* | 600866 | 69.48 | 0.00 |
| | ***TBP*** | 600075 | 6.48 | 0.02 |
| | ***PSMB1*** | 602017 | 33.38 | 0.97 |
| | *FAM120B* | 612266 | 80.03 | 0.00 |
| | ***DLL1*** | 606582 | 4.65 | 1.00 |
| | *ERMARD (C6orf70)* | 615532 | 84.86 | 0.00 |
| | *DYNLT2* | 186977 | - | 0.00 |
| | *PHF10* | 613069 | 18.13 | 0.00 |
| | *C6orf120* | 616987 | 92.18 | - |
| | *THBS2* | 188061 | 59.96 | 0.56 |
| | *SMOC2* | 607223 | 55.41 | 0.00 |
| | *DACT2* | 608966 | 93.50 | 0.01 |
| | *KIF25* | 603815 | 95.70 | 0.00 |
| | ***AFDN*** | 159559 | - | 1.00 |
| | *TCP10L3* | 187020 | - | 0.00 |
| | *UNC93A* | 607995 | 85.97 | 0.00 |
| | *GPR31* | 602043 | 91.50 | 0.02 |
| | *CCR6* | 601835 | 84.13 | 0.01 |
| | *CEP43* | 605392 | - | 0.00 |
| | *RNASET2* | 612944 | 88.27 | 0.00 |
| | *RPS6KA2* | 601685 | 64.44 | 0.02 |
| | *MPC1* | 614738 | 49.44 | 0.04 |
| | *TBXT* | 601397 | - | 0.06 |
| | ***PDE10A*** | 610652 | 52.14 | 1.00 |
| 6q26 | ***QKI*** | 609590 | 4.10 | 0.75 |
| | *CAHM* | 615930 | - | - |
| | *PACRG* | 608427 | 10.61 | 0.00 |
| | ***PRKN*** | 602544 | 0.77 | 0.00 |
| | *AGPAT4* | 614795 | 61.98 | 0.10 |
| | ***MAP3K4*** | 602425 | 59.19 | 1.00 |
| | *PLG* | 173350 | 40.11 | 0.01 |
| | *LPA* | 152200 | 78.22 | 0.00 |
| 6q25.3 | *LPAL2* | 611682 | - | - |
| | *SLC22A3* | 604842 | 49.06 | 0.00 |
| | *SLC22A2* | 602608 | 63.53 | 0.00 |
| | *SLC22A1* | 602607 | 75.70 | 0.00 |
| | *AIRN* | 604893 | - | - |
| | ***IGF2R*** | 147280 | 53.96 | 1.00 |
| | *MAS1* | 165180 | 28.09 | 0.00 |
| | *MRPL18* | 611831 | 49.78 | 0.00 |
| | ***TCP1*** | 186980 | 5.10 | 1.00 |
| | *ACAT2* | 100678 | 53.11 | 0.00 |
| | ***WTAP*** | 605442 | 6.76 | 1.00 |
| | ***SOD2*** | 147460 | 0.99 | 0.15 |
| | *FNDC1* | 609991 | 75.28 | 0.00 |
| | *TAGAP* | 609667 | 82.32 | 0.85 |

| | | | | |
|---|---|---|---|---|
| | *RSPH3* | 615876 | 86.62 | 0.00 |
| | ***EZR*** | 123900 | 8.64 | 0.18 |
| | *DYNLT1* | 601554 | 66.63 | 0.00 |
| | *TMEM181* | 613209 | 67.29 | 0.01 |
| | *GTF2H5* | 608780 | 30.21 | 0.05 |
| | *SERAC1* | 614725 | 61.19 | 0.00 |
| | *SYNJ2* | 609410 | 63.42 | 0.00 |
| | *SNX9* | 605952 | 40.36 | 0.71 |
| | *ZDHHC14* | 619295 | 29.53 | 0.72 |
| | ***ARID1B*** | 614556 | 14.17 | 1.00 |
| | *NOX3* | 607105 | 38.77 | 0.00 |
| | *TFB1M* | 607033 | 53.74 | 0.00 |
| 6q25.2 | *TIAM2* | 604709 | 53.43 | 0.68 |
| | ***SCAF8*** | 616024 | 25.56 | 1.00 |

All OMIM genes studied, extending from 6q27 to 6q25.2 (16 Mb). Predicted HI-genes are highlighted in bold. The HI and pLI scores were derived from https://decipher.sanger.ac.uk in May 2021
